# Serum extracellular vesicles trace COVID-19 progression and immune responses

**DOI:** 10.1101/2022.01.19.22269529

**Authors:** Kevin Ho Wai Yim, Simone Borgoni, Richard Chahwan

**Affiliations:** Institute of Experimental Immunology, University of Zurich, Zurich, Switzerland

**Author notes:** **Corresponding author:** Richard Chahwan.

**Keywords:** COVID-19, SARS-CoV-2, extracellular vesicles, diagnosis, clinical liquid biopsy

## Abstract

Coronavirus disease 2019 (COVID-19) has transformed very quickly into a world pandemic with severe and unexpected consequences on human health. Concerted efforts to generate better diagnostic and prognostic tools have been ongoing. Research, thus far, has primarily focused on the virus itself or the direct immune response to it. Here, we propose extracellular vesicles (EVs) from serum liquid biopsies as a new and unique modality to unify diagnostic and prognostic tools for COVID-19 analyses. EVs are a novel player in intercellular signaling particularly influencing immune responses. We herein show that innate and adaptive immune EVs profiling, together with SARS-CoV-2 Spike S1^+^ EVs provide a novel signature for COVID-19 infection. It also provides a unique ability to trace the co-existence of viral and host cell signatures to monitor affected tissues and severity of the disease progression. And provide a phenotypic insight into COVID-associated EVs.

## INTRODUCTION

Coronavirus disease 2019 (COVID-19) was first identified in December 2019 in Wuhan, China. The disease progressed into a global pandemic with over 64 million confirmed cases and over 3.9 million confirmed deaths as of June 2021^[1]^. COVID-19 patients can be asymptomatic, suffer from mild symptoms such as fever, cough, and dyspnea or develop into severe conditions characterized as acute respiratory distress syndrome (ARDS) requiring mechanical ventilation^[2]^. SARS-CoV-2, a positive-sense, single-stranded RNA virus, is known as the causative pathogen of COVID-19. The commonly acknowledged mechanism of SARS-CoV-2 pathology is the entry of viruses into angiotensin-converting enzyme 2 (ACE2) expressing host cells, with a tropism for different organs, such as the respiratory tract, kidneys, liver, heart, brain, and blood vessels^[3]^. SARS-CoV-2 infected cells can recruit different immune cell types and induce innate inflammatory responses as well as adaptive immune responses mediated by targeted antibodies. Sars-CoV-2 specific immunoglobulins (Ig) types M, A, and G have been used as an indicator of protective immunity in infected patients. However, such antibody responses normally emerge around 10 to 21 days after infection and may take even longer (four weeks or more) in mild cases to be detected^[4]^. In general, around 5 % of COVID-19 patients develop severe conditions like ARDS, which arises around one week after symptom onset. Therefore, SARS-CoV-2 specific antibody titer measurement is not the best predictor of severe disease for infected patients who show mild symptoms early after infection but rapidly develop ARDS. Extracellular vesicles (EVs) are nanosized lipid-bilayer vesicles that carry nucleic acid and protein cargo. They are constitutively secreted by virtually all types of cells and circulate in most biofluids such as blood, urine, saliva, and breast milk. Since the surface markers and molecular cargo of EVs can reflect the cellular origin and activation status, they have been utilized as non-invasive biomarkers from liquid biopsies in the past decade, for diagnostic and prognostic purposes^[5–7]^. Thanks to the lipid-bilayer structure of EVs, they are intrinsically more stable than naked circulating molecules such as antibodies and cytokines, conferring a higher potential to provide a more robust and long-lasting effect on the host immune response. Here, we characterized serum EVs from healthy donors, early COVID-19 patients (< 13 days from symptom onset), and late COVID-19 patients with mild disease (> 13 days from symptom onset), in terms of size distribution, concentration and surface marker profile using nanoparticle flow analyzer (NanoFCM^®^) ^[8]^. Cluster analysis of different EVs subpopulations based on surface marker expression was performed to identify signatures of healthy donors, early COVID-19 and late COVID-19 patients. Lastly, COVID-19 specific SARS-CoV-2 Spike S1^+^ serum EVs were characterized in relation to disease progression and host immune responses to determine disease severity.

## RESULTS

### Multiplex profiling of serum EVs derived from mild COVID-19 patients

To explore the landscape of serum immune EVs during SARS-CoV-2 infection, we sampled a cohort of 20 mild COVID-19 patients and 17 healthy donors (Table 1). According to the WHO definition (World Health Organization, 2020), all the sampled COVID-19 patients in this study experienced mild illness with symptoms such as fever, fatigue, or dyspnea. Serum EVs were isolated from the whole blood of donors and patients. Purified serum EVs were analyzed by nanoparticle analyzer to examine the size distribution and concentration of different serum EVs subsets with a dedicated panel of immune markers and tetraspanins marker (Fig. 1A).

**Table 1.** Demographic and clinical characteristics of the patient cohort.

|  | Healthy | Mild COVID-19 |  |
| --- | --- | --- | --- |
| No. of samples | 17 | 20 |  |
| Median age [years] | 31 | 34 |  |
| Gender (M/F) | 7/10 | 11/9 |  |
| Time since symptom onset (days) | - | 11 ± 5.01996 |  |
| **Laboratories values** |  |  | **P value** |
| Hemoglobin(mean ± SD, [g/l]) | 143.13 ± 11.76 | 81.35 ± 75.79 | 0.0028 |
| Absolute platelet count(mean ± SD, [G/l]) | 252 ± 50.71 | 243.25 ± 61.79 | ns |
| Total white blood cell count mean ± SD, [G/l]) | 6.06 ± 1.73 | 5.13 ± 1.14 | ns |
| Monocytes(mean ± SD, [G/l]) | 0.45 ± 0.16 | 0.46 ± 0.1 | ns |
| Neutrophils(mean ± SD, [G/l]) | 3.61 ± 1.29 | 2.64 ± 0.91 | 0.0129 |
| Eosinophils(mean ± SD, [G/l]) | 0.1 ± 0.05 | 0.12 ± 0.09 | ns |
| Basophils(mean ± SD, [G/l]) | 0.05 ± 0.01 | 0.03 ± 0.01 | 0.0015 |
| Lymphocytes(mean ± SD, [G/l]) | 1.84 ± 0.56 | 1.85 ± 0.56 | ns |
| CD3- CD56bright CD16dim NK cells<br>(mean ± SD, [cells/ul]) | 18.47 ± 5.35 | 12.1 ± 5.7 | ns |
| CD3- CD56dim CD16bright NK cells<br>(mean ± SD, [cells/ul]) | 164.59 ± 119.87 | 248.25 ± 131.75 | ns |
| CD4+ T cells(mean ± SD, [cells/ul]) | 376.24 ± 468.03 | 780.3 ± 290.92 | 0.0029 |
| CD19+ B cells(mean ± SD, [cells/ul]) | 101.41 ± 130.44 | 189.4 ± 103.36 | 0.0282 |
| C-reactive protein(mean ± SD, [mg/l]) | 1.04 ± 0.97 | 1.74 ± 1.89 | ns |
| LDH(mean ± SD, [U/l]) | 332.24 ± 53.88 | 342.39 ± 79.04 | ns |
| IL-6(mean ± SD, [pg/ml]) | 0.47 ± 0.57 | 2.51 ± 4.24 | ns |
| IL-10 (mean ± SD, [pg/ml]) | 1.15 ± 1.3 | 1.64 ± 2.19 | ns |
| IFNγ (mean ± SD, [pg/ml]) | 0.88 ± 1.68 | 1.89 ± 2.64 | ns |
| TNFα(mean ± SD, [pg/ml]) | 7.32 ± 3.04 | 8.36 ± 3.49 | ns |
| Anti-CoV-2 IgA(mean ± SD, [μg/mL]) | 0.34 ± 0.18 | 4.85 ± 7.2 | 0.0144 |
| Anti-CoV-2 IgG(mean ± SD, [μg/mL]) | 0.27 ± 0.15 | 1.03 ± 0.92 | 0.0019 |
| **Comorbidities** |  |  |  |
| Hypertonia - no. (%) | - | - |  |
| Diabetes - no. (%) | - | - |  |
| Heart disease - no. (%) | - | 1 (5%) |  |
| Lung disease - no. (%) | - | - |  |
| Malignancy - no. (%) | - | - |  |
| Immunosuppression - no. (%) | - | - |  |
| Kidney disease - no. (%) | - | - |  |
| Cerebrovascular disease - no. (%) | - | - |  |
| M Crohn - no. (%) | - | 1 (5%) |  |
| Allergic asthma - no. (%) | - | 2 (10%) |  |
| Hypothyreose - no. (%) | 1 (6%) | 3 (15%) |  |

**Figure 1.**
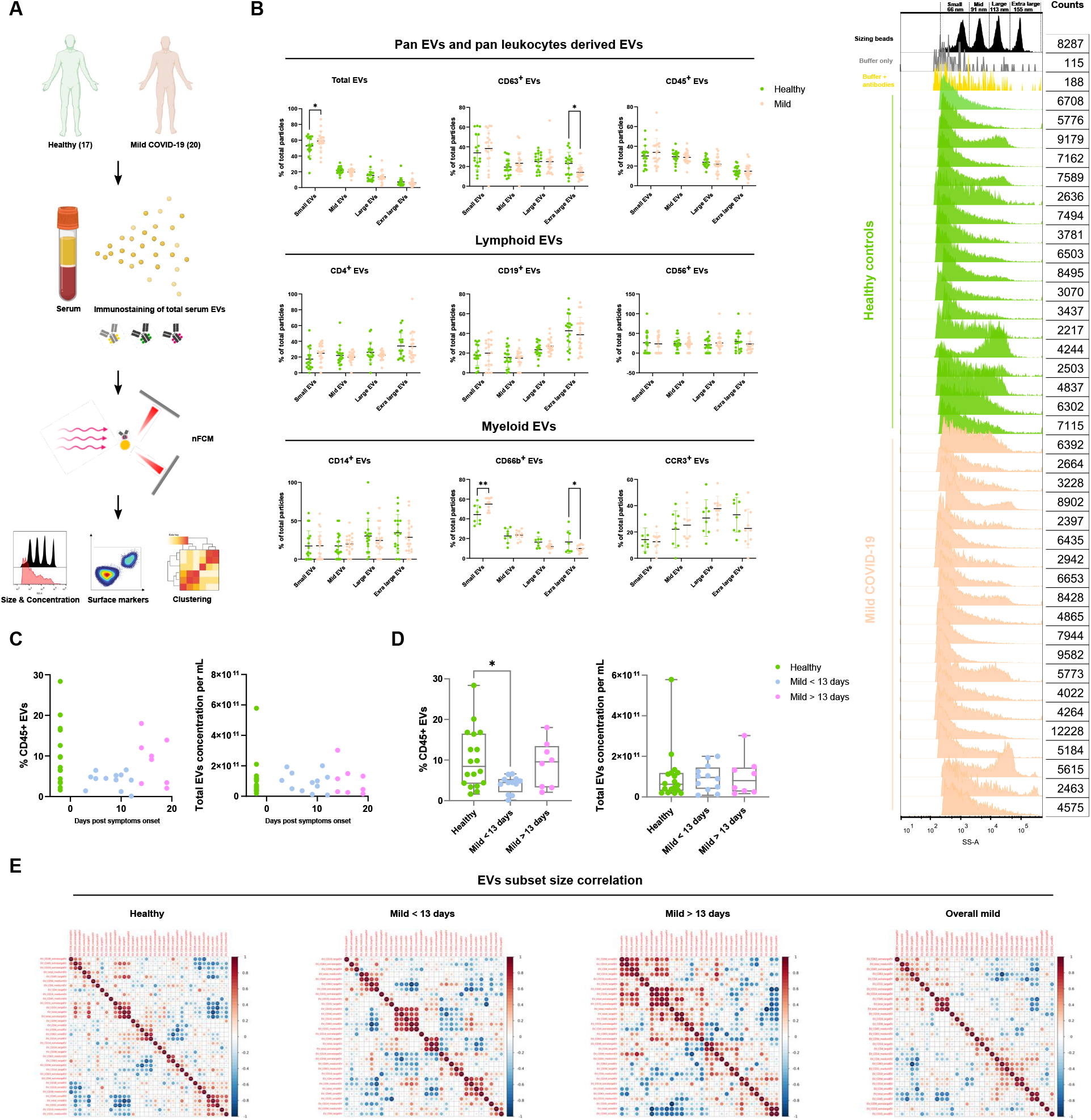
Characterization of immune serum EVs in healthy controls and mild COVID-19 patients. (A) Schematic outline of EVs profiling from denoted human samples. (B) Size distribution quantification of serum EVs from denoted human samples and different EV subsets, with size reference beads with a mixture of four modal sizes of 66 nm (small), 91 nm (medium), 113 nm (large), 155 nm (extra-large). Representative side scatter histogram of size reference beads in (B) and total serum EVs from denoted human samples on the right. (C, D) Quantification of total serum EVs and CD45+ EVs in denoted human samples at days of reported symptom onset. (E) Spearman’s rank correlation matrix of size distribution of serum EVs subsets between healthy donors and mild COVID-19 patients. One-way ANOVA, p < 0.05 *, p < 0.01 **, p < 0.005 ***.

According to MISEV 2018 guideline, the physical properties of EVs, such as particle size distribution and concentration were analyzed to ensure the reproducibility of the result. Moreover, for diagnostic interest, potential variance in particle size distribution of different serum EVs subsets between healthy controls and COVID-19 patients might provide valuable predictive information (Fig. S1a). Using a mixture of four different sizes (66, 91, 113, 155 nm) of monodisperse silica nanoparticles (refractive index = 1.461), we applied 4 size interval bins based on the 4 separated side-scatter burst areas to quantify the approximate size distribution of serum EVs, abbreviated as small, medium, large, extra-large accordingly (Fig. S1b). In COVID-19 patients small EVs were more enriched, and CD66b^+^ EVs showed an increase compared to healthy controls (p < 0.05, p < 0.01 respectively). However, extra-large EVs were reduced in CD63^+^, CD38^+^, IgA^+^, IgG^+^ EVs in COVID-19 patients compared to healthy controls (p < 0.05) (Fig. 1B, C, S1d).

Immune cells profiling of COVID-19 patients revealed numerous alterations in both innate and adaptive immunity. However, whether immune cells derived EVs were influenced by COVID-19 or involved in any form of disease specific responses remains unknown. Canonical leukocyte marker CD45 was first examined in patient serum EVs to gain an overview of immune EVs changes in COVID-19. Interestingly, CD45^+^ serum EVs showed a significant reduction in mild COVID-19 patients compared to healthy control (Fig. S1c). We next visualized the alteration of CD45^+^ EVs levels against days post symptoms onset. Significant depletion in CD45^+^ EVs in patients between 3 to 13 days post symptoms onset was observed compared to healthy controls (p < 0.05). Strikingly, patients after 13 days post symptoms onset displayed a recovery of CD45^+^ EVs level comparable to healthy controls. This finding highlighted the importance to dissect the analysis into pre- and post-13 days post symptoms onset to gain more precise perspectives in the serum EVs dynamics in COVID-19. Interestingly, depletion of CD45^+^ EVs in the early onset of COVID-19 correlated to CD45^+^ cells deficiency observed in severe COVID-19 patients reported by another study ^[9]^, indicating the high sensitivity and early detection capacity of EVs based diagnostics. Total EVs concentration did not show significant differences suggesting the alteration in CD45^+^ EVs level was independent of total EVs production during SARS-CoV-2 infection (Fig. 1C, D). The correlation between sizes and markers in EVs subsets between healthy controls and COVID-19 patients was visualized by Spearman’s rank correlation matrix (Fig. 1E). A significant correlation of large and extra-large CD31^+^ EVs (predominantly expressed by endothelial tissues) and total EVs was observed in healthy controls and post-13 days patients but not in pre-13 days patients. There was also a strong correlation between the small CD14^+^, CD19^+^, CD56^+^ and CD63^+^ EVs in pre-13 days patients which were not present in healthy controls nor post-13 days patients. These data suggest that classical monocytes-, B cells- and natural killer cells-derived small EVs are predominantly affected in the early stage of COVID-19.

### Detection of SARS-CoV-2-Spike S1^+^ serum EVs

ACE2 containing EVs have been reported to prevent infection by SARS-CoV-2 virus indicating the relevance of EVs in COVID-19 progression^[10]^. Moreover, SARS-CoV-2 Spike S1 containing EVs have been shown to serve as decoys for neutralizing antibodies^[11]^. To explore whether SARS-CoV-2 utilizes such strategies to influence disease progression and interaction with the host, we attempted to detect and profile Spike S1^+^ serum EVs in this cohort of patients. To eliminate the possibility of virus contamination in serum EVs purification, viral RNA detection by PCR was performed with patients’ serum and none of the specimens tested positive. Specificity of Spike S1 antibody has been validated with negative control HEK293A and Spike S1 transfected HEK293A and their released EVs, in combination with serial titration of staining cocktail with recombinant Spike S1 proteins (Fig. S2).

Spike S1^+^ EVs were detected in a small subset of healthy controls, suggesting their serum EVs might be carrying cross-reactive epitopes that bind to Spike S1 antibodies used in the study. We, therefore, applied a cut-off of Spike S1 fluorescence signal at 1.75% of total particles to exclude all healthy controls. 5 out of 16 mild COVID-19 patients (dark blue dots in plots) had > 1.75% Spike S1^+^ serum EVs (classified as Spike S1^+^ EVs positive), suggesting the existence of SARS-CoV-2 specific EVs in human serum which could subsequently influence disease progression (Fig. 2A). To further identify the origin of Spike S1^+^ EVs, we co-stained Spike S1 with a panel of immune and endothelial markers since SARS-CoV-2 are known to significantly affect endothelial tissues in patients^[12]^. Notably, Spike S1^+^CD31^+^ EVs levels were significantly increased in pre-13 days COVID-19 group compared to healthy controls and post-13 days COVID-19, but not with the immune markers analyzed (Fig. S3). Elevated levels of endothelial markers including CD31 were reported in COVID-19^[13]^, and temporary stress triggers CD31^+^ microparticle release^[14]^. These data suggest Spike S1^+^ EVs likely originate from SARS-CoV-2 infected endothelial tissues. Total CD31^+^ EVs level also showed an elevating tendency in COVID-19 patients compared to healthy controls. Reduction of small Spike S1^+^ EVs was also observed in post-13 days patients compared to pre-13 days patients (Fig. 2B).

**Figure 2.**
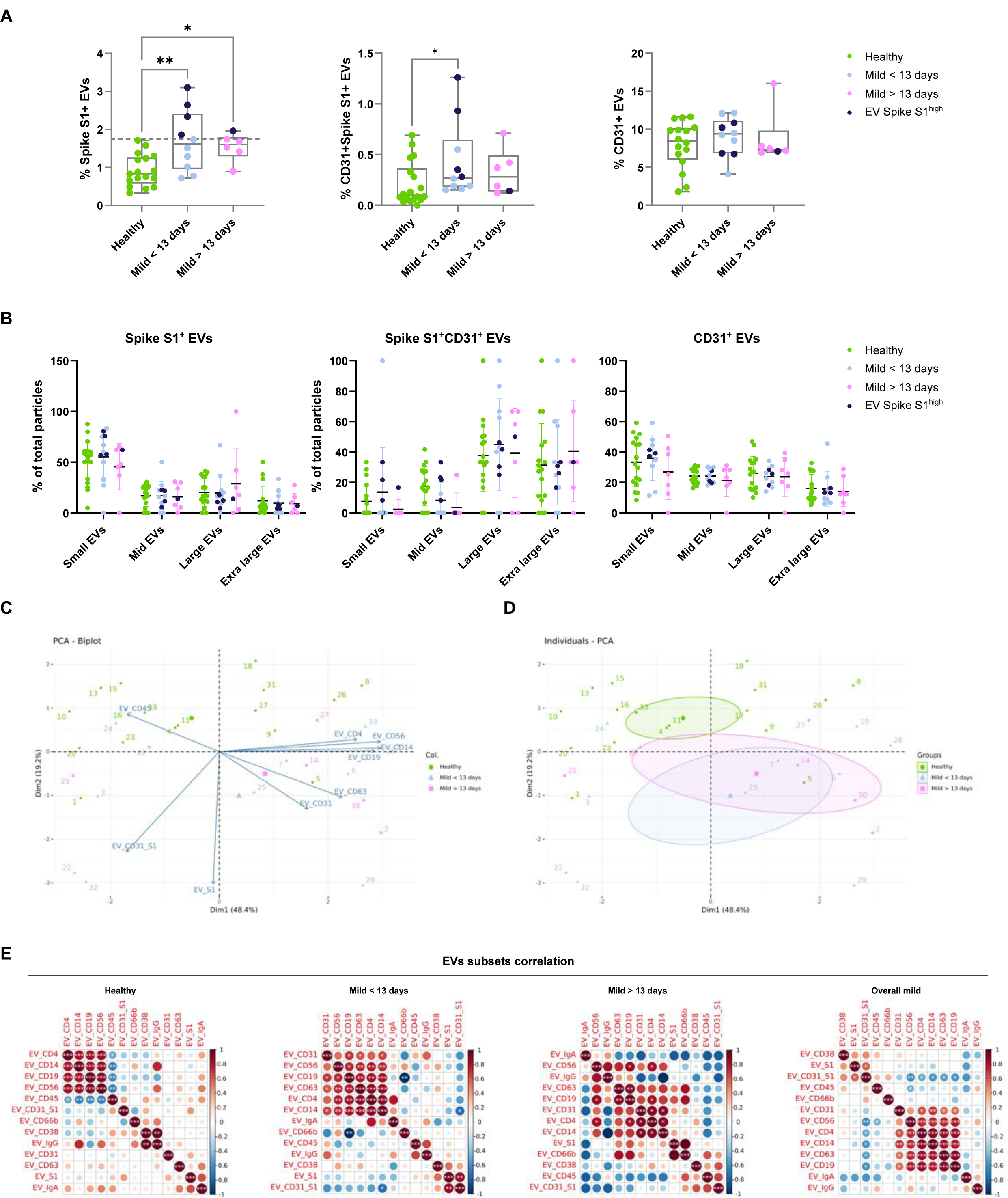
Characterization of Sars-Cov-2 Spike S1+ serum EVs in healthy controls and mild COVID-19 patients. (A) Quantification of Spike S1+, Spike S1+CD31+ and CD31+ serum EVs in denoted human samples, dark blue (Spike S1^+^ EVs positive). (B) Size distribution quantification of serum EVs from denoted human samples and indicated EV subsets. (C) Bi-plot and principal component analysis of set of serum EVs markers defining healthy and mild COVID-19 status. (D) Spearman’s rank correlation matrix of serum EVs subsets between healthy donors and mild COVID-19 patients. One-way ANOVA, p < 0.05 *, p < 0.01 **, p < 0.005 ***.

To gain deeper insights into the relationship between serum EVs and disease status, we applied principal component analysis of serum EVs subset level across the cohort (Fig. 2C). A biplot revealed the contribution of different EVs subsets as arrows showing the significant association of CD45^+^ EVs and healthy controls. A quartile was dominated by pre-13 days patients and highly associated with Spike S1^+^ EVs. A quartile with mostly post-13 days was defined by high levels of CD31^+^ and CD63^+^ EVs. In another quartile, a heterogeneous mix of predominantly healthy controls was defined by high levels of CD4^+^, CD14^+^, CD19^+^ and CD56^+^ EVs. Individuals PCA clustering allowed the stratification of disease status based on frequencies of different EVs subsets across the cohort. Taken together, despite the anticipated diversity across individuals and less pronounced phenotypes in mild COVID-19 compared to severe ones, these exploratory analyses enable the usage of serum EVs to predict disease status on top of canonical clinical approaches (Fig. 2D).

To further probe the alterations of different serum EVs subsets during COVID-19, we applied hierarchical clustering in a correlation map of serum EVs subsets across the cohort (Fig. 2E). In the healthy controls group, specialized immune cell derived EVs, CD4^+^, CD14^+^, CD19^+^ and CD56^+^ EVs had a strong positive correlation with each other but negatively correlate with CD45^+^ EVs, suggesting, in healthy state, immune EVs are less likely to originate from lymphocytes nor monocytes but other types of leukocytes (i.e., neutrophils, around 60% of leukocytes in blood). However, such negative correlation did not persist in COVID-19 patients, indicating the surge of lymphocytes and monocytes driven responses during SARS-CoV-2 infection. Significant positive correlation was also observed between CD38^+^ and IgG^+^ EVs in healthy controls but not in any COVID-19 group. In the pre-13 days mild COVID-19 group, CD31^+^ and CD63^+^ EVs started to positively correlate with specialized immune EVs. Strong positive correlation of Spike S1^+^ and Spike S1^+^CD31^+^ EVs and negative correlation of CD19^+^ and CD66b^+^ EVs were also observed. In the post-13 days mild COVID-19 group, the strong positive correlation between specialized immune EVs was reduced as well as the correlation of Spike S1+ and Spike S1+CD31+ EVs. In the overall mild COVID-19 group, regardless of time after symptoms onset, specialized immune EVs positively correlate with CD31^+^ EVs and Spike S1^+^ EVs also positively correlate with Spike S1+CD31+ EVs. In summary, the analysis of serum EVs cluster signatures in relation to disease status and time after symptoms onset uncovered the dynamic patterns of serum EVs subset levels during COVID-19 progression.

### Abundance of SARS-CoV-2-Spike S1 serum EVs indicate host immunological responses

To better understand the dynamics of Spike S1^+^ EVs during SARS-CoV-2 infection, we visualized the alteration of Spike S1^+^ and Spike S1^+^CD31^+^ EVs across the cohort against time after symptoms onset (Fig. 3A). Spike S1^+^ EVs positive patients were present mostly in pre-13 days post symptoms onset, two of which from day 10 showed high Spike S1^+^CD31^+^ EVs (Fig. 3A). To explore the immunological relevance of serum Spike S1^+^ EVs during COVID-19 progression, we analyzed the SARS-CoV-2 specific immunoglobulins levels across the cohort and in relation to time after symptoms onset (Fig. 3B). SARS-CoV-2 specific IgA and IgG levels were below 0.6 G/l in all healthy controls and showed an increasing trend close to 10 days post symptom onset (Fig. 3B, 3C). Two of the Spike S1^+^ EVs positive patients at day 10 showed elevated levels of SARS-CoV-2 specific IgA and IgG (up to 24 and 3 G/l respectively) which are predominantly observed in severe COVID-19 cases, suggesting the dynamics Spike S1^+^ EVs levels in patients could provide sensitive detection of alternated immunological responses.

**Figure 3.**
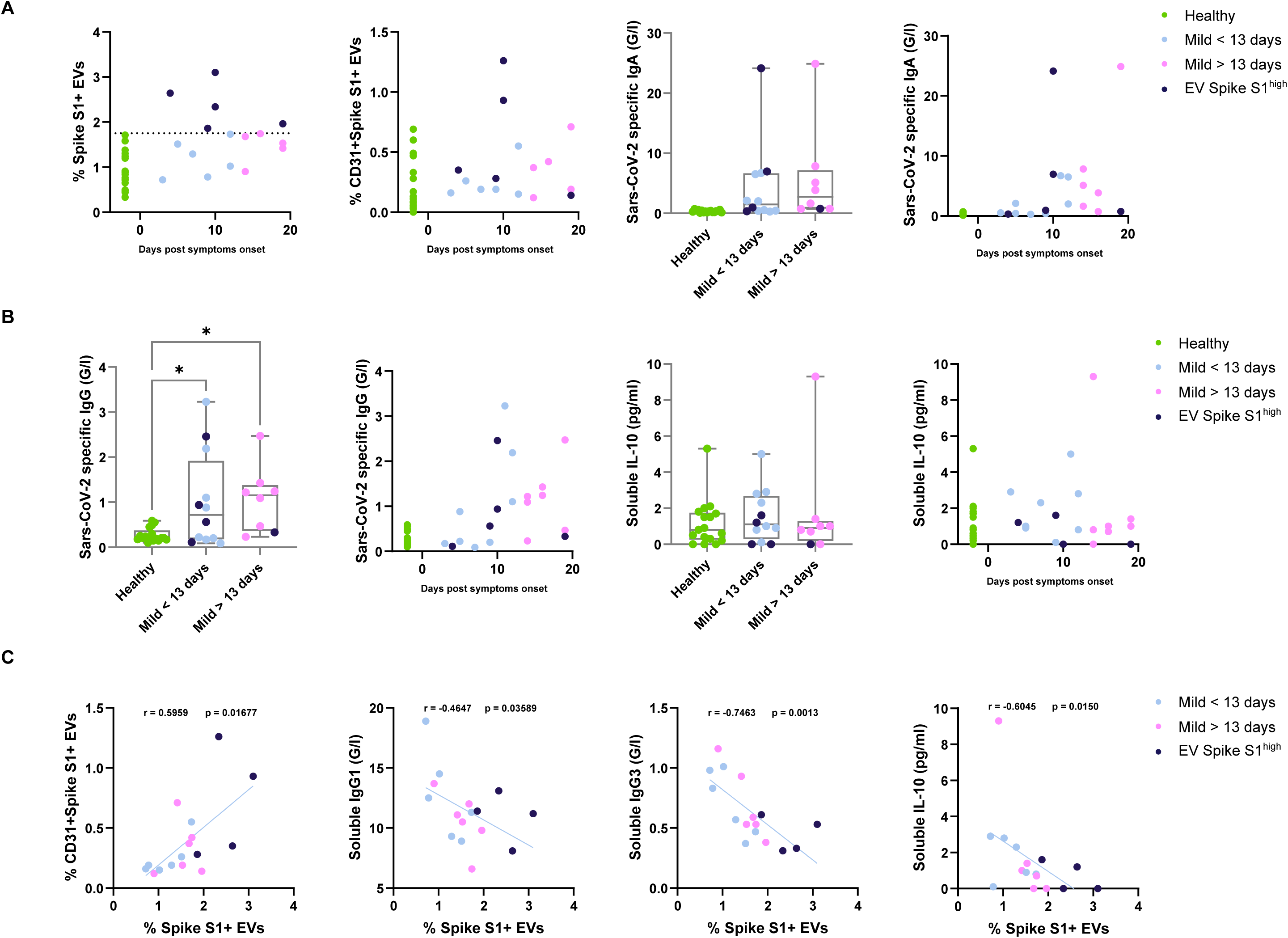
Correlation of Sars-Cov-2 Spike S1+ serum EVs with host immune responses in healthy controls and mild COVID-19 patients. (A) Quantification of Sars-Cov-2 Spike S1+ and Sars-Cov-2 Spike S1+CD31+ EVs in denoted human samples at days after reported symptom onset. (B, C) Quantification of soluble Sars-Cov-2 specific immunoglobulins in denoted human samples at days after reported symptom onset. (D) Quantification of soluble interleukin-10 levels in denoted human samples at days after reported symptom onset. (E) Spearman’s correlation of level of Spike S1+ serum EVs to Spike S1+CD31+ EVs, soluble IgG1, IgG3 and interleukin-10 levels.

Levels of immunosuppressive cytokine IL-10^[15]^, one of the hallmarks observed in severe COVID-19 was compared across the cohort. IL-10 levels were comparable between healthy controls and COVID-19 patients, indicating cytokine secretion was not significantly affected in this cohort of mild COVID-19 patients (Fig. 3D).

To further understand the relationship between Spike S1^+^ EVs and other significantly altered factors between healthy controls and COVID-19 patients, we performed direct correlation analysis between the levels of Spike S1^+^ EVs and Spike S1^+^CD31^+^ EVs, viral specific immunoglobulins (IgG1 and IgG3) and immunosuppressive IL-10 (Fig. 3E). A strong positive correlation was observed between Spike S1^+^ EVs and Spike S1^+^CD31^+^ EVs (r = 0.6, p = 0.017), suggesting the Spike S1^+^ EVs are likely to originate from endothelial tissues. We also found a strong negative correlation between Spike S1^+^ EVs and IgG1 (r = -0.46, p = 0.036) and IgG3 (r = -0.75, p = 0.0013). In contrast to SARS-CoV-2 specific IgA and IgG, IgG1 and IgG3 are predominantly induced by viral infection, with IgG3 appearing first during infection^[16]^. These negative correlations indicate levels of Spike S1^+^ EVs could estimate whether the host antibody mediated responses are SARS-CoV-2 specific. Another negative correlation was observed between Spike S1^+^ EVs and immunosuppressive IL-10, suggesting patients with higher Spike S1^+^ EVs are less prone to immunosuppression of the immune system and possibly a lower chance of disease deterioration.

### Mild COVID-19 patients derived serum EVs affect healthy PBMCs responses ex vivo

Immune cells derived soluble factors such as IFN-_γ_, IL6, and TNF are known to cause inflammation in severe COVID-19 cases and subsequently cause complications in the disease. Since EVs have been shown to modulate immune responses in viral infection, we sought to explore the possibility of EVs mediated immune regulation in COVID-19. B cells mediated antibodies neutralization and T cells mediated cytokines production are the key drivers of host immune defenses against COVID-19, understanding B and T cells responses in the presence of purified serum EVs from the cohort would enable us to better understand the immune regulatory network in COVID-19. To understand such mechanism, we activated healthy PBMC CD19^+^ B cells (with IL-4 and IL-21) and CD3^+^ T cells (anti-CD3/CD28 and PMA/Ionomycin) *ex vivo* in the presence of PBS control and purified serum EVs across the cohort. Cell viability, activation, antibodies class switching (in B cells), and cytokine production (in T cells) were measured 4 days after activation by flow cytometry (Fig. 4A).

**Figure 4.**
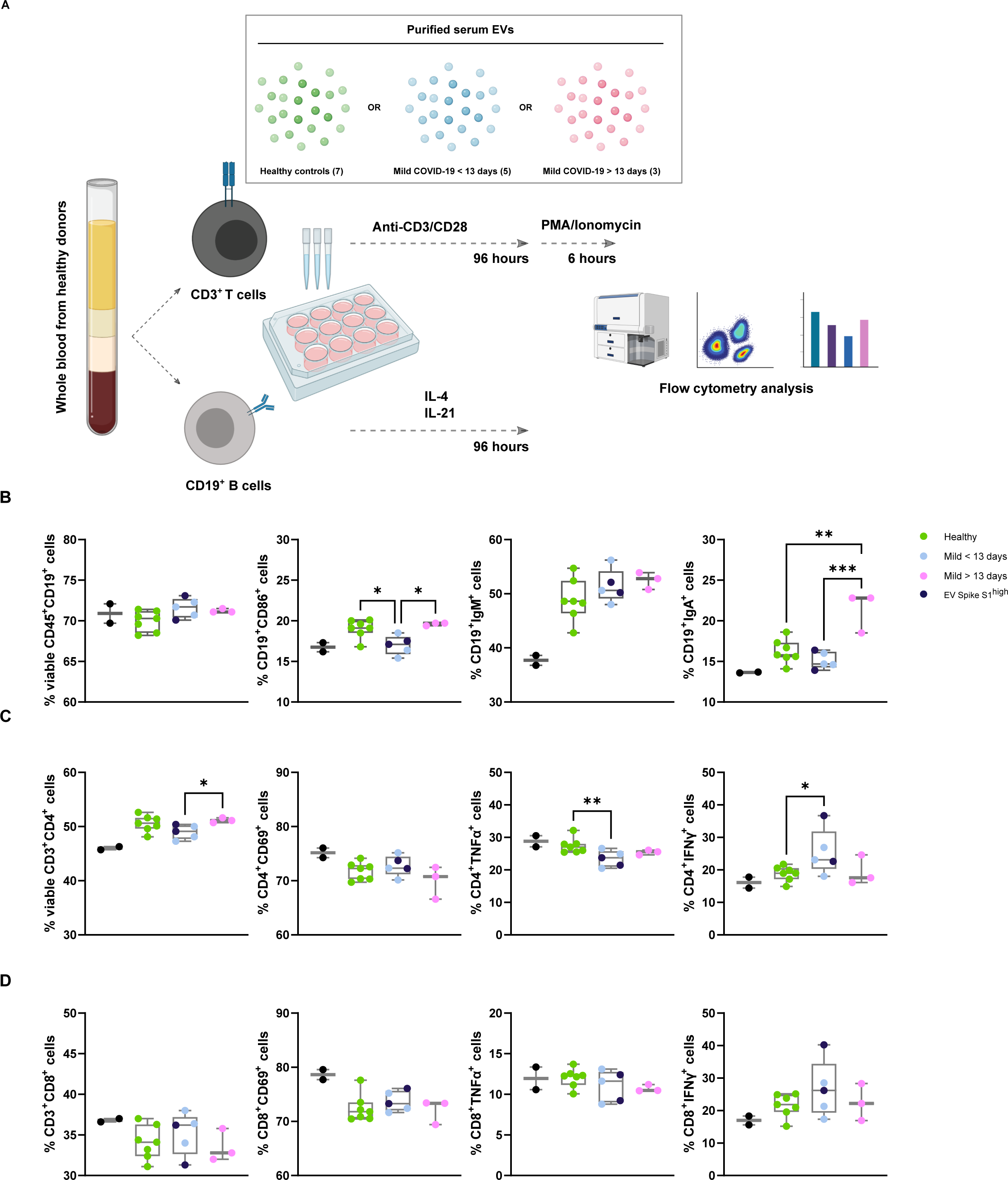
Mild COVID-19 patients derived serum EVs affect healthy PBMCs responses ex vivo (A) Schematic outline of *ex vivo* healthy PBMCs activation in the presence of PBS control and serum EVs from healthy donors and mild COVID-19 patients. (B-D) Quantification of different subsets expansion, activation, cytokine production (T cells) (C, D) and class switch recombination (B cells) (B). One-way ANOVA, p < 0.05 *, p < 0.01 **, p < 0.005 ***.

Viability of CD19^+^ B cells was consistent across the serum EVs in the cohort. Activation marker CD86 expression was reduced in pre-13 days COVID-19 derived EVs compared to both healthy controls and post-13 days COVID-19 derived EVs, suggesting serum EVs from pre-13 days suppress B cells activation. IgM expression was similar across the serum EVs in the cohort while IgA expression was significantly increased in post-13 days COVID-19 EVs, implying post-13 days COVID-19 EVs promote antibodies class switching.

Viability of CD4^+^ T cells was reduced in pre-13 days COVID-19 derived EVs compared to post-13 days COVID-19 derived EVs but not observed in CD8^+^ T cells. Activation marker CD69 expression in CD4^+^ and CD8^+^ T cells was similar across the serum EVs in the cohort. Significant reduction of TNF production in CD4^+^ T cells was observed in pre-13 days COVID-19 EVs, with a reducing tendency in post-13 days COVID-19 EVs compared to healthy controls. However, TNF production in CD8^+^ T cells was comparable across the serum EVs in the cohort, suggesting CD4^+^ T cells are more susceptible to serum EVs modulation. IFN-_γ_ production of CD4^+^ and CD8^+^ T cells was increased in pre-13 days COVID-19 EVs compared to both healthy controls and post-13 days COVID-19 derived EVs. Taken together, serum EVs from COVID-19 patients are capable to influence B and T cells responses in a targeted manner that possibly contribute to the immune phenotypes observed in COVID-19 patients (Fig. 4B).

## DISCUSSION

Throughout the COVID-19 pandemic, immune cell-profiling studies continue showing a wide spectrum of immunological complications from mild to severe cases such as lymphopenia, hyper inflammation and series of cytokine storms. Aside from studied soluble immune factors like antibodies and cytokines, EVs are emerging as novel and potent intercellular signaling mediators. EV cargos are under study as predictive tools for COVID-19 severity^[17–19]^. However, such recent studies required mass spectrometry and RNA-seq of EVs cargo which is not easy to convert into routine clinical diagnostics due to the tedious processing and high expertise required. Or they studied covid related EVs *ex vivo* ^[11]^. We focus here on i) serum immune-derived EVs because immune cells are predominantly found in the bloodstream, as are their released EVs; ii) viral-specific EVs to gain specificity and viral tracking ability; iii) on analyzing early- and late-mild covid patients to study the effect of the virus with minimal symptom complications; and iv) attributing phenotypic functions for COVID-19 associated EVs. Therefore, our analysis has consequences that surpass the basic investigation of COVID-19 biomarkers per se and instead tries to trace the dynamic response of the virus and our body’s response to it.

To harness the full potential of immune EVs in COVID-19, we applied multi-parametric EVs phenotyping to characterize total EVs and specific immune associated subsets. The high sensitivity and resolution of NanoFCM^®^ used in this study allowed the possibility of direct single EVs phenotyping. We could simultaneously measure and correlate EV size, concentration and marker subset quantification; surpassing the conventional antibody coated beads approach currently used^[20]^. General enrichment of small EVs (∼ 66 nm) in mild COVID-19 patients was observed and more evident in total serum EVs and CD66b^+^, CD38^+^, IgA^+^, IgG^+^ EVs. Interestingly, such enrichment of small EVs seems to correlate with reduction in extra-large EVs (∼ 155 nm) and is more pronounced in CD63^+^ EVs and CD66b^+^ EVs. Immunological studies have shown inflammatory phenotypes in granulocytes (expressing CD66b) are one of the strongest discriminators between non-infected and infected individuals as well as between severity status of COVID-19 patients^[21]^. The size shifts we observed correspond to the moderate enrichment of smaller EVs due to cellular stress reported by others^[22]^. Our results suggest that EV’s biogenesis machinery in general especially granulocytes may have been altered by SARS-CoV-2 infection causing cellular stress. Although the underlying mechanism of such phenotype is yet to be determined, it is conceivable that this is partly due to the interaction between SARS-CoV-2 and the endosomal sorting complexes required for EV transport^[23]^. Moreover, size distribution correlation maps revealed a strong positive association between small specialized immune cells derived EVs (CD14^+^, CD19^+^, CD56^+^; classical monocytes, B cells, NK cells) in pre-13 days after symptoms onset, suggesting these immune compartments were potentially more stressed in the early phase of COVID-19. Another highlight in our study is the depletion of leukocyte-derived CD45^+^ EVs in early phase of mild COVID-19 (pre-13 days). Although our EV data is supported by studies measuring CD45 cell counts^[9]^, we also observed a recovery of CD45^+^ EV levels in the latter phase of mild COVID-19 (post-13 days). Given the predominant 14 days COVID-19 isolation guidelines^[24]^, and the fact that these patients were all mildly symptomatic our data bolsters the ability of our study to dynamically trace covid19 progression based on immune EVs. This also may reflect the partial recovery of host immune systems as reflected by serum EVs markers. It is worth noting that by combining pre- and post-13 days data these small dynamic immune changes can counter-weigh each other and be lost to measurements, which suggest that future work should consider infection time or disease onset as a factor. Moreover, biplot cluster analysis showed insignificant variance of EVs subsets distribution between age groups and sex of donors (Fig. S4).

Engineered EVs expressing SARS-CoV-2 antigens, such as ACE2 and Spike S1, have been shown to perform potent functions in inhibiting coronavirus infection and serving as decoys for neutralizing antibodies, respectively^[10,11]^. These findings not only prove the potential relevance of EVs in COVID-19 progression but also highlight the possibility of SARS-CoV-2 utilizing EVs as an intermediate driver for replication and interplay with host immune defenses. Within our cohort of mild COVID-19 patients, almost one-third showed significantly elevated Spike S1^+^ EVs level in our platform when compared to healthy controls. The existence of Spike S1^+^ EVs in patients’ serum – which we show for the first time and which seems to peak 8-12 days post symptoms – strongly support the potential of SARS-CoV-2 exploiting host EVs to act as decoys for neutralizing antibodies and evade from the host immune surveillance and specific responses as anticipated in an *in vitro* study of artificial Spike S1^+^ EVs^[11]^. Moreover, the possibility to detect SARS-CoV-2 Spike S1 marker from patients’ serum EVs and profile their dynamics during course of infection provided a novel tool for disease specific EVs phenotyping in both research and clinical applications, especially in monitoring life-threatening severe COVID-19 progression. Throughout our panel of immune markers and endothelial marker co-stained with Spike S1 across the cohort, Spike S1^+^CD31^+^ EVs levels were significantly altered in pre-13 days COVID-19 compared to healthy controls. These data support the notion that endothelial tissues (predominantly expressing CD31) were significantly influenced by SARS-CoV-2 infection, but also suggesting a notable amount of Spike S1^+^ EVs originate from endothelial tissues. Further studies with staining of additional markers as well as future development on high dimensional single EVs analysis shall provide a more conclusive representation of serum EVs subsets distribution not only in COVID-19, but also other viral infections and diseases.

Within our dataset, a global PCA analysis of analyzed EVs subsets distribution across the cohort displayed a clear stratification between healthy controls, pre-, and post-13 days of COVID-19. The partial overlap between pre-13 and post-13 days revealed the core serum EVs features associated with the SARS-CoV-2 infection and might be explained by the ‘long COVID’ observed in other follow-up studies^[25,26]^. Despite the overlap, post-13 days COVID-19 were less separated from pre-13 days COVID-19 indicating the gradual normalization of analyzed serum EVs, possibly associated with their parental cells and tissues. From the correlation clustering of EVs subsets across the cohort, we could show distinct serum EVs signatures that enable differentiation between healthy controls and COVID-19 patients as well as tracing the progression of COVID-19.

Furthermore, correlation of Spike S1^+^ EVs and SARS-CoV-2 specific IgA, IgG, IL-10, IgG1 and IgG3 revealed the clinical implications of Spike S1^+^ EVs levels in the context of host immune responses and disease progression. Within our dataset, Spike S1^+^ EVs positive patients tend to show higher SARS-CoV-2 specific antibody responses and lower levels of immunosuppressive IL-10, which in theory, should result in less disease deterioration. Negative correlation between Spike S1^+^ EVs and generic viral induced IgG1 and IgG3 responses^[16]^ implies that Spike S1^+^ EVs levels could indicate the chances of developing SARS-CoV-2 specific responses in an early predictive manner, and subsequently, enable more precise and prompt treatment decision in the clinics.

Our functional assay of purified serum EVs in *ex vivo* PBMC B and T cells activation demonstrated novel perspectives of the biological relevance and influences of COVID-19 derived serum EVs. The significant increase in IgA^+^ B cells in the presence of post-13 days COVID-19 EVs indicates certain EV subsets derived from later phase of COVID-19, are partially facilitating B cell antibody class switching during COVID-19 progression. Our dataset also aligns with the elevated IFN-_γ_ levels observed in severe COVID-19 patients^[27]^, IFN-_γ_ enrichment in CD4^+^ T cells in the presence of pre-13 days COVID-19 EVs but not post-13 days COVID-19 EVs. This dataset suggests that early in SARS-CoV-2 infection, infected host cells were possibly reprogrammed to produce certain EVs subsets to promote inflammatory IFN-_γ_ production in CD4^+^ T cells. Interestingly, despite TNF elevation were observed in other clinical studies of severe COVID-19 patients^[28]^, we found significant depletion of TNF in CD4^+^ T cells in the presence of pre-13 days COVID-19 EVs, suggesting serum EVs from early phase of COVID-19 are suppressive for TNF production in CD4^+^ T cells. Taken together, our findings provide an initial stepping-stone for the field to understand EVs signaling mediated COVID-19 progression.

With the rather small cohort size and limited markers combinations, the predictive values of serum EVs could be further bolstered and fully exploited in clinical applications. Future work shall consider increasing the size of cohort with the addition of severe patients from various group cohorts including age, sex, genetic, geographical background, and treatment courses. Moreover, to enhance the diagnostic and prognostic specificity of serum EVs in COVID-19, future work shall include sampling from non-SARS-CoV-2 infected individuals such as seasonal influenza patients. This approach will allow us to better stratify the EVs subsets signatures between COVID-19 and general flu-like viral infections and ultimately fortify clinical predictive value of EVs. Indeed, future optimization of high-dimensional spectral analyzers for EVs characterization with the addition of more immune and viral markers will broaden our spectrum to better trace the dynamics of host immune responses as well as disease progression. In the therapeutic context, our functional assay demonstrated the feasibility of utilizing serum EVs from different disease severity to manipulate host immune responses. Nevertheless, identification of responsible EVs subsets and understanding their mode of action are essential for us to harvest their therapeutic potentials in the future.

The dynamics of serum EVs subsets distribution highlighted their predictive values in the perspectives of overall host immune responses and correlation between disease specific Spike S1^+^ EVs and immune responses during COVID-19 progression. The study strengthens the potential of serum EVs based diagnostic and prognostic, potentially therapeutic applications in COVID-19, and easily transferred to other types of viral infections and cancers.

## MATERIALS AND METHODS

### Study subjects

Peripheral blood was draw from healthy donors and COVID-19 patients recruited at the University Hospital Zurich (Switzerland) outpatient clinic. The patients were eligible if they were symptomatic at the time of inclusion, had a newly diagnosed SARS-CoV-2 infection confirmed by quantitative reverse-transcriptase polymerase chain reaction (RT-qPCR), and were over 18 years old. Healthy donors (n = 17) were recruited as controls. All participants, patients and healthy controls, signed a written informed consent. This non-interventional, observational study was approved by the Cantonal Ethics Committee of Zurich (BASEC #2016-01440) and performed in accordance with the Declaration of Helsinki. The sample size was based on availability of the samples. Investigators were blinded to disease severity, while performing experiments. While the analysis was cross-sectional, the patient outcomes were recorded prospectively after inclusion. Standard clinical laboratory data (CRP, LDH, complete blood count with differential) was collected from the first day of hospitalization. Patients were classified according to WHO guidance into mild cases (n=20) in the same hospital laboratory. All healthy controls were tested for SARS-CoV-2 specific IgA and IgG antibodies, and all were below the diagnostic reference value. All patients received a standard clinical laboratory sampling and cytokines were measured. Furthermore, samples from 20 mild COVID-19 patients and all healthy subjects were processed for nano flow analysis and *ex vivo activation of PBMCs*. Briefly, 3 – 5 mL of peripheral venous blood was collected from into BD Vacutainer serum clot activator 10 ml (367896). After collection, tubes were left vertically undisturbed on the bench for 15 minutes to allow blood clot and followed by centrifugation at 2,500 g for 10 mins at 4°C for separation of sera. The supernatant was collected into a new tube and the serum samples were stored at 4°C until use. Due to the limited number of available samples, certain serum samples did not have enough amount for all the immunostaining panels.

### Isolation of serum EVs for phenotyping analysis and functional assays

1 mL of serum samples were first diluted with 9 mL of PBS and concentrated using Amicon® ultra-0.5 centrifugal filter devices (Millipore, Amicon® Ultra 100 K device) at 3,000 g for 30 mins at 4°C. Serum retentate (100 uL per patient) was diluted with 1.4 mL of PBS and subjected to centrifugation at 10,000 g for 30 mins at 4°C. The 10k pellet was resuspended in 1.5 mL PBS and subjected ultracentrifugation at 120,000 g for 90 mins at 4°C (F50L-24×1.5 rotor). The pellet was resuspended in 50 uL PBS and stored at 4°C until use.

### Nano flow analysis of serum EVs

The configuration NanoFCM^®^ used in the study was performed as described in another study^[20]^. 10 uL of purified serum EVs were subjected to immunofluorescence staining using antibodies listed in supplementary materials (Table 1) at the concentration of 1 in 200 uL for 1 hour at 4°C covered in dark. Immuno-stained serum EVs were washed by ultracentrifugation at 120,000 g for 90 mins at 4°C, pellet was resuspended in 50 uL PBS for nano flow analysis. Monodisperse silica nanoparticles of four different sizes, with modal peak sizes of 66 nm, 91 nm, 113 nm and 155 nm were used as the size reference standard to calibrate the size distribution of EVs.

### Ex vivo activation of PBMCs

Buffy coats were obtained from Blutspende Zurich, Schlieren, Switzerland (BASEC #2019-00837). PBMCs were isolated using Ficoll-Paque density gradient centrifugation. CD19^+^ B cells were isolated using STEMCELL EasySep™ Release Human CD19 Positive Selection Kit according to manufacturer’s instruction. CD19^+^ B cells were activated with both human recombinant IL-4 and IL-21 at 10 ng/mL (Biolegend). Remaining cell suspension were plated in RPMI-1640 media supplemented in 10% heat inactivated fetal bovine serum (FBS) (Sigma) and 1% penicillin streptomycin/glutamine (Thermo) for 24 hours at 37°C, 5% CO_2_. Cells were collected at 300 g for 5 mins and stimulated with anti-CD3/CD28 beads at 1 bead per cell (Thermo). 50 uL of purified serum EVs from healthy controls, pre-13 days and post-13 days mild COVID-19 were added to cultured cells together with stimulatory agents.

### Cell cultures transfection and flow cytometry for SARS-CoV-2 Spike S1 staining controls

HEK293A cells were grown in 10 cm tissue culture dishes in DMEM (Sigma) supplemented with 10% heat-inactivated FBS and 1% penicillin/streptomycin. To generate Spike S1 expressing HEK293A cells and EVs, we transfected HEK293A cells with pCMV14-3X-Flag-SARS-CoV-2 S plasmid (Addgene #145780) together with GFP plasmid (for transfection efficiency quantification) and EVs were collected 24 hours post transfection from conditioned medium using serial centrifugation as noted above. Transfection efficiency of HEK293A was measured by flow cytometry (BD Canto II) in the FITC channel. To test the antibody binding specificity, prior to immunostaining of Spike S1 on cells and EVs, molar ratios of 0:1, 1:1, 2:1, 5:1 of recombinant Spike S1 proteins (Sino Biological, #40591-V08H) were incubated with anti-Spike S1 for 30 mins at 4°C to neutralize unbound antibodies. Resulting staining cocktail (with range of neutralizing recombinant Spike S1 proteins) were used to stain non-transfected and Spike S1 transfected HEK293A and EVs for 1 hour at 4°C covered in dark. Spike S1 signals were detected by flow cytometry (HEK293A) and nano flow analyzer (EVs).

### Data and statistical analysis

Both cells and EVs flow cytometry data were exported as FCS files and analyzed using Flowjo software. Statistical analysis of clinical diagnostic and flow cytometry values were performed using Graphpad (version 9.1.1, GraphPad Software, La Jolla California USA). Ordinary One-way ANOVA, P-value < 0.05 was considered as statistical significance. PCA and biplot were generated with R software (version 4.0.1) using the package ‘factoextra’ and ‘ggplot2’. Spearman’s correlation analyses were produced using the corrplot package and implemented hclust method. The correlations between different EVs subsets, and between different disease severity group were analyzed using non-parametric Spearman correlations. The significance threshold was set at alpha < 0.05.

## Data Availability

All data produced in the present work are contained in the manuscript

## ACKNOWLEDGMENT

We sincerely thank Prof Onur Boyman and Dr Carlo Cervia for providing critical elements for this study including clinical samples, data, and insightful comments. We thank Dr Ben Peacock for NanoFCM sample support. RC is supported by BBSRC (BB/N017773/2), SNF (CRSK-3_190550; IZSEZ0_204655), and the UZH Research Priority Program (URPP Translational Cancer Research). KY is supported by a Bio & MedTech Entrepreneur Fellowship.

## AUTHOR CONTRIBUTIONS

KY and RC conceived the idea and coordinated the project. RC secured funding and guided the work. KY performed experiments and analyses. KY and SB performed in vitro experiments and statistical data analyses. KY wrote the manuscript draft, which was then finalized by KY and RC with the assistance of SB.

## SUPPLEMENTARY FIGURE LEGENDS

**Figure S1**. Nanoflow control and experimental setup. (A) Representative nanoflow gating strategies of EVs, HEK293A derived EVs (control) and representative patient derived serum EVs. (B) Representative back gating strategies to determine size distribution of EV subsets using size reference beads with mixture of four modal sizes, 66 nm (small), 91 nm (medium), 113 nm (large), 155 nm (extra-large). (C) Quantification of CD45^+^ serum EVs in healthy controls and mild COVID-19 patients. (D) Quantification of size distribution of CD38^+^, IgA^+^, IgG^+^ serum EVs in healthy controls and mild COVID-19 patients.

**Figure S2**. Binding specificity of Sars-CoV-2 Spike S1 antibodies. (A) Representative flow gating strategies of HEK293A co-transfected with GFP and Spike S1 plasmid after 24 hours. (B) Competition of anti-Spike S1 binding in HEK293A in (A) with addition of recombinant Spike S1 proteins in denoted molar ratio. (C) Representative flow gating strategies of EVs derived from HEK293A co-transfected with GFP and Spike S1 plasmid after 24 hours and competition of anti-Spike S1 binding in EVs with addition of recombinant Spike S1 proteins in denoted molar ratio.

**Figure S3**. (A) Representative flow gating strategies of patient derived serum EVs for Spike S1 and CD31. (B) Quantification of Spike S1^+^CD45^+^, Spike S1^+^CD38^+^, Spike S1^+^CD56^+^, Spike S1^+^IgA^+^, Spike S1^+^IgG^+^, Spike S1^+^CD66b^+^ serum EVs in in healthy controls and mild COVID-19 patients.

**Figure S4**. PCA plot clustering of serum EVs samples based on age (A) and sex (B).

