## Supplementary figures and images for "Serum extracellular vesicles trace COVID-19 progression and immune responses"

### Figure S1

#### Supplementary figure 1

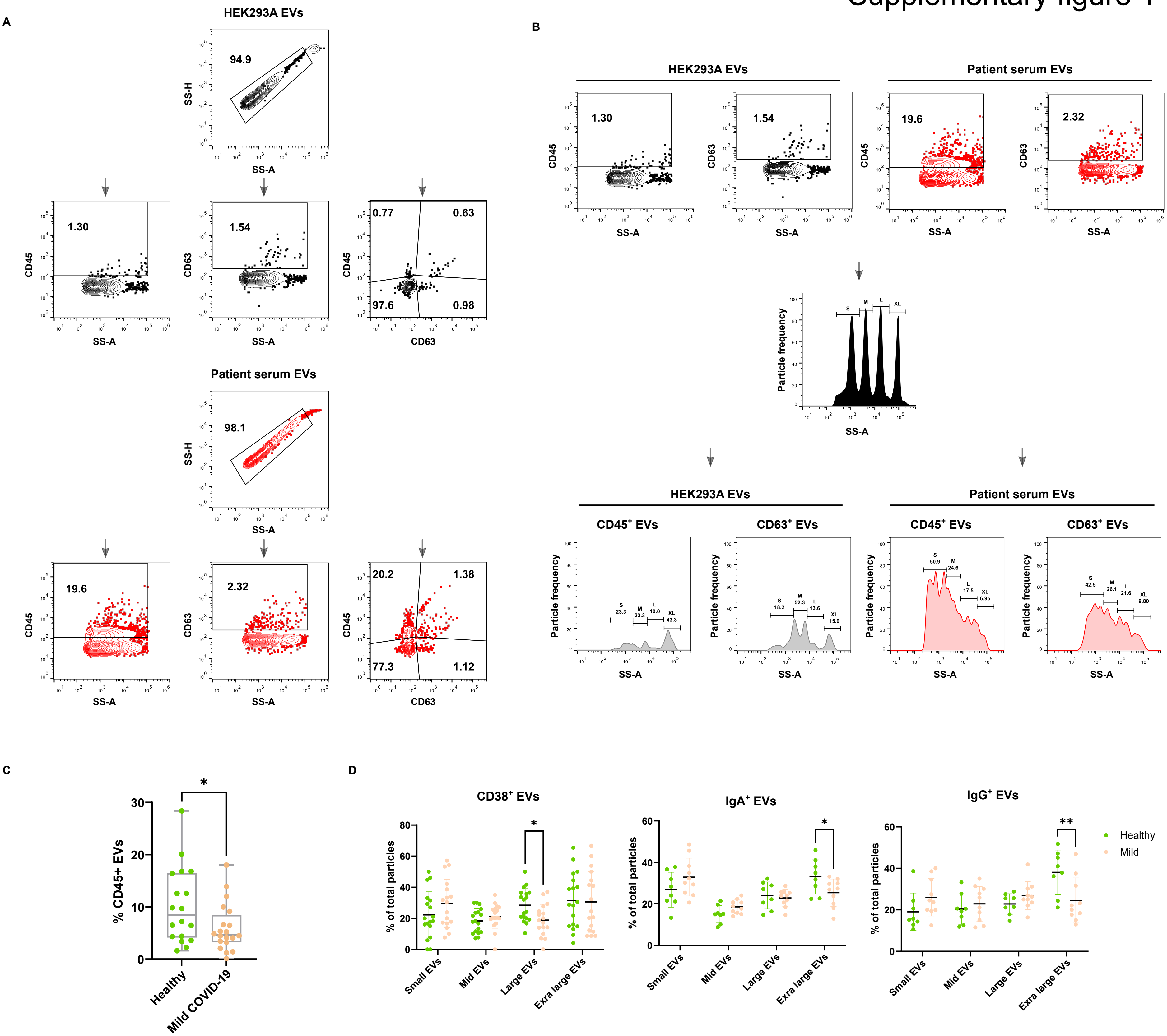

### Figure S2

#### Supplementary figure 2

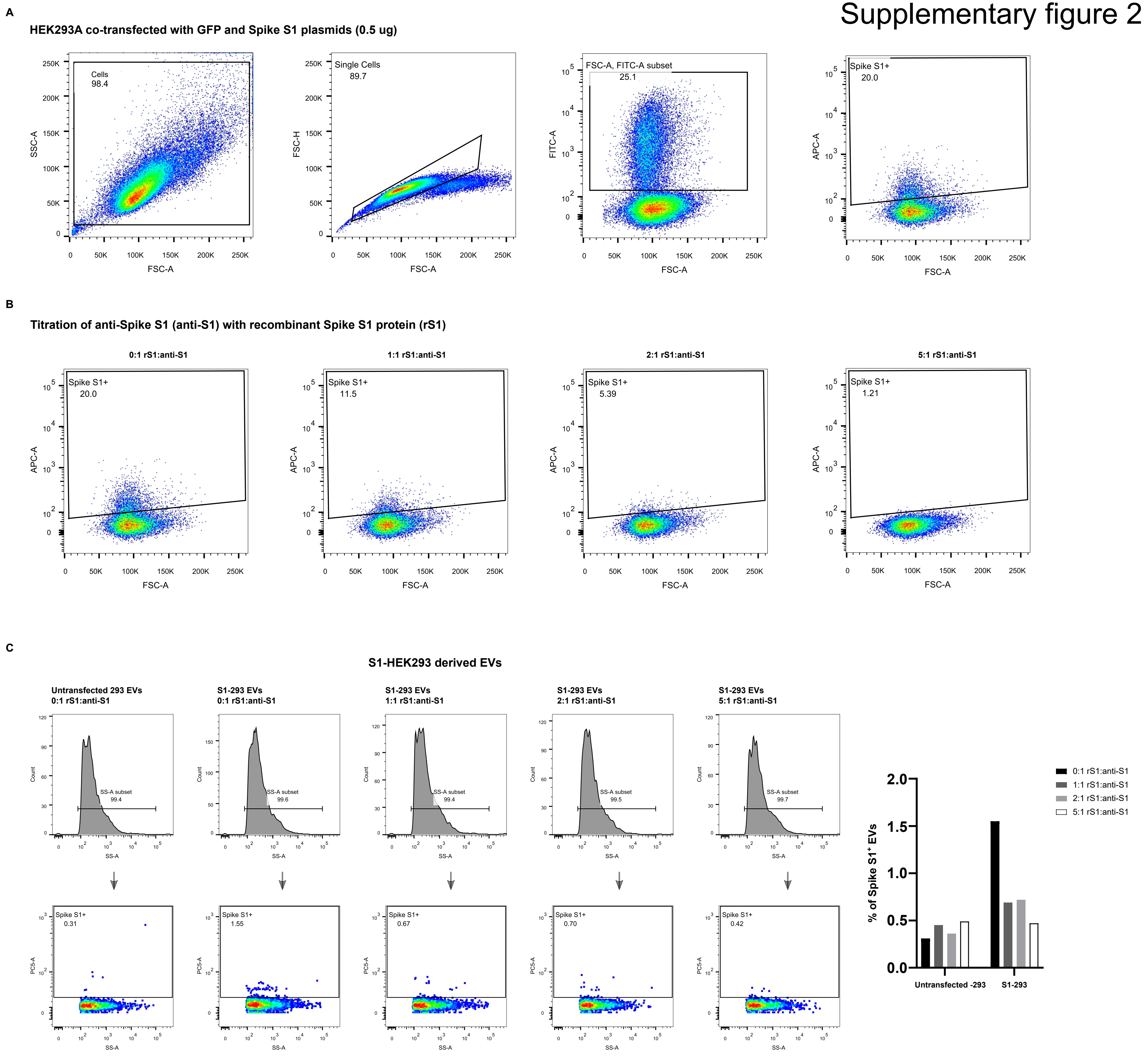

### Figure S3

Supplementary figure 3

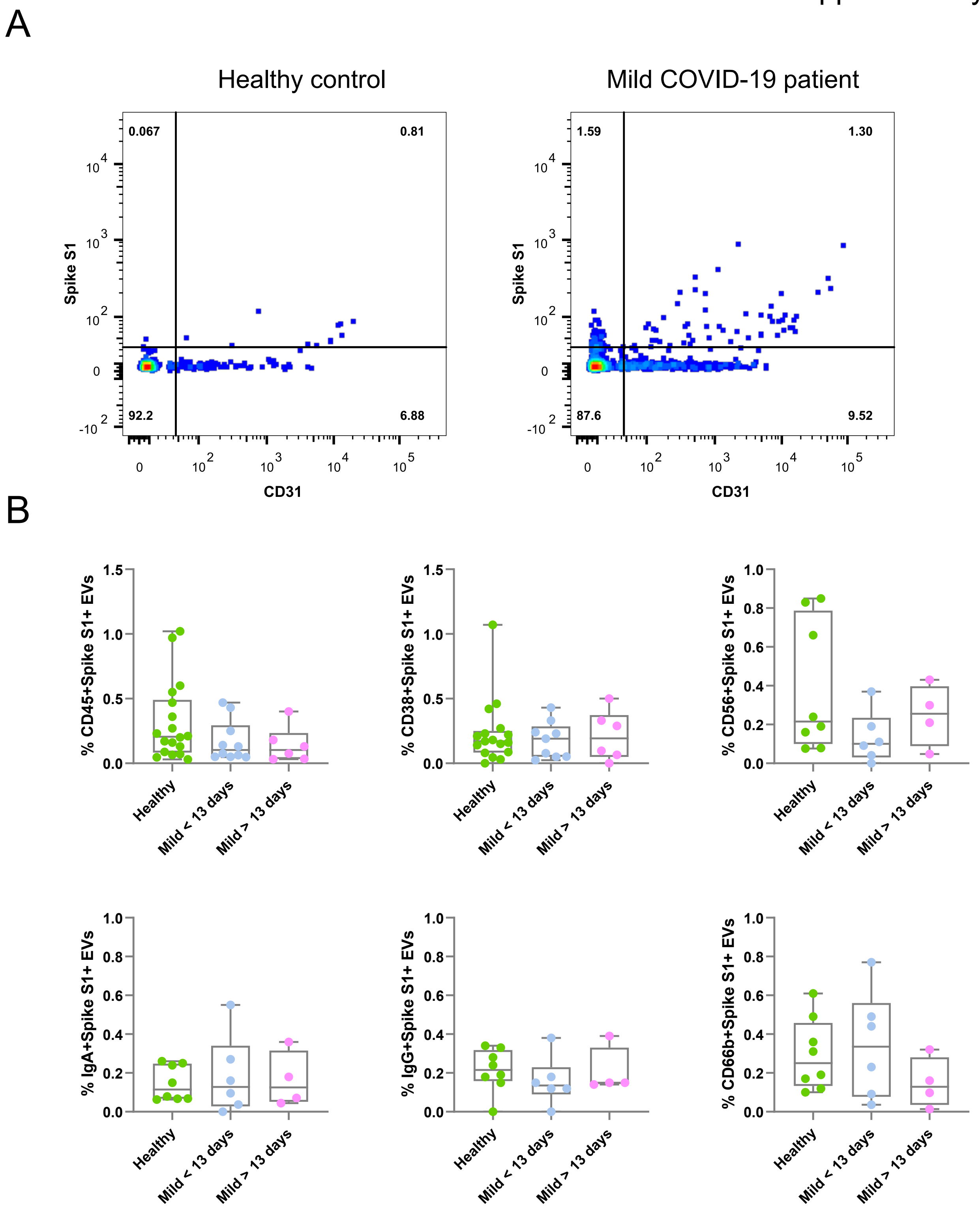

### Figure S4

#### Supplementary figure 4

**A**

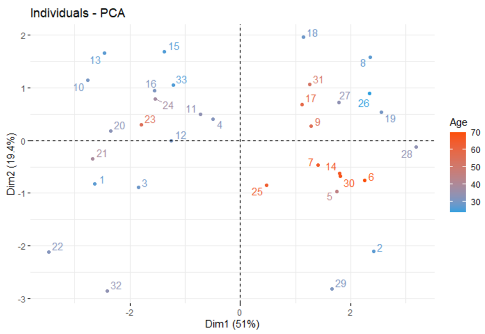


**B**

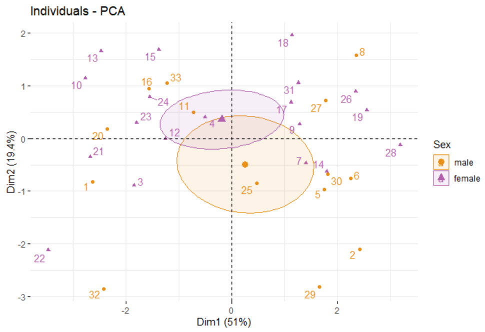
